# Risk Factors for Clinically Relevant Postoperative Pancreatic Fistula (CR-POPF) after Distal Pancreatectomy: A Single Center Experience

**DOI:** 10.1101/2020.05.04.20090241

**Authors:** Gao Qing Wang, Dipesh Kumar Yadav, Wei Jiang, Yong Fei Hua, Cai De Lu

**Author notes:** Indicates equally contributed in study design, data collection, data analysis, and preperation of the manuscript (Gao Qing Wang, Dipesh Kumar Yadav). Corresponding author: Cai De Lu, Department of Hepatobiliary and Pancreatic Surgery Li HuiLi Hospital Ningbo 315040 Zhejiang, China.

## Abstract

Clinically relevant postoperative pancreatic fistula (CR-POPF) is the considerable contributor to major complications after pancreatectomy. The purpose of this study was to evaluate the potential risk factor contributing to CR-POPF following distal pancreatectomy (DP) and discussed the risk factors of pancreatic fistula in order to interpret the clinical importance. All the patients who underwent DP in between January 2011 and January 2020 were reviewed retrospectively in accordance with relevant guidelines and regulations. The univariate and multivariate analysis was performed was performed to test an independent risk factors for pancreatic fistula. P<0.05 was considered statistically significant. In all of the 263 patients with DP, pancreatic fistula was the most common surgical complication 19.0%. The univariate analysis of 18 factors showed that the patients with a malignant tumor, soft pancreas, and patient without ligation of the main pancreatic duct are more likely to develop pancreatic fistula. However, on multivariate analysis the soft texture of the pancreas (OR= 2.381, P= 0.001) and the ligation of main pancreatic duct (OR= 0.388, P= 0.002) were only an independent influencing factor for CR-POPF. As a conclusion, pancreatic fistula was the most common surgical complication after DP, and the texture of pancreas and ligation of main pancreatic duct can influence an incidence of CR-POPF.

## Introduction

In recent decades, distal pancreatectomy (DP) has become a common surgical technique for the treatment of benign and malignant pancreatic tumors, chronic pancreatitis and pancreatic trauma^1^. Technically, DP is a simpler procedure compared to pancreaticoduodenectomy (PD), as a pancreato-enteric anastomosis is seldom required, and prevention of postoperative pancreatic fistula remains a challenge in DP due to an ineffective closure of the pancreatic remnant. The incidence of pancreatic fistula after DP ranges from 5% to 32%, depending upon the definition used and the underlying pancreatic pathology^2-6^. As per updated definition of the International Study Group for Pancreatic Fistula (ISGPF) in 2016, a postoperative pancreatic fistula (POPF) is an external fistula with a drain output of any measurable volume of fluid after postoperative day 3 with an amylase level more than 3 times the upper limit, associated with a clinically relevant condition (i.e Grade B and Grade C). Additionally, clinical criteria must be met in order to be considered as true pancreatic fistula, earlier Grade A postoperative pancreatic fistula is now no longer considered as true pancreatic fistula, instead its now reported as a “biochemical leak”^7^.

Clinically relevant postoperative pancreatic fistula (CR-POPF) is the considerable contributor to major complications such as bleeding, abdominal abscess, sepsis, and even death following pancreatic resection^8-10^. Nevertheless, various attempts has been made to improve surgical outcomes that includes suture closure of the pancreatic stump, staple transection of the pancreas, the use of fibrin glue to cover the pancreatic stump, coverage of the pancreatic stump with autologous tissue, the use of pancreatic stents, and the use of prophylactic octreotide^10^. Woefully, most of these methods have failed to improved fistula rates^10, 11^. Nonetheless, risk identification and risk stratification might benefit in the prevention of POPF. Indeed, the development of the Fistula Risk Score (FRS) for PD and its application has provided a great understanding for the prediction of POPF and has guided the of modern accessible mitigation techniques in reduction of morbidity^10, 12^. However, the underlying mechanism of POPF after DP is still poorly understood and FRS for DP has not been developed yet that can predict the risk of POPF.

The purpose of this study was to evaluate the potential risk factors contributing to CR-POPF following DP and discussed the risk factors of pancreatic fistula in order to interpret the clinical importance.

## Patients and Methods

### Patients

All the patients who underwent DP at the Li HuiLi Hospital, Ningbo between January 2011 and January 2020 were reviewed retrospectively from Electronic Medical Record System and were approved by an institutional review board of the Li HuiLi Hospital. Written informed consent were obtained from the patients or patients party and was consistent with the Declaration of Helsinki^13^. The present data analysis includes 263 patients (n= 213 underwent open distal pancreatectomy and n= 50 cases underwent laparoscopic distal pancreatectomy) undergoing DP over a 8-year period. Data were collected from the medical records on the on standardized data sheets for all patients and the variables collected were patients demographics, surgery indications, preoperative evaluation and risk evaluation, preoperative lab values, perioperative, and postoperative course, that includes, age, body mass index (BMI), smoking, preoperative American Society of Anesthesiologists (ASA) risk grading^14^, indication for surgery, pancreas texture, combined multivisceral resection, splenectomy, ligation of main pancreatic duct, pancreatic stump treatment, preoperative diabetes, intraoperative blood loss, use of somatostatin after surgery, preoperative albumin level, postoperative albumin level (3 days after surgery), surgical approach (open vs laparascopic), operation time, and pancreatic resection range.

### Treatment Protocols

All patients underwent preoperative contrast-enhanced abdominal computed tomography, or enhanced magnetic resonance imaging examination with cholangiopancreaticography (MRCP) to accurately assess the nature of the lesion, location, size and the relationship with the splenic vessels and other organs. Additionally, perioperative prophylactic antibiotics and a daily dose of low molecular weight heparin (LMWH) was given to all patients. Moreover, all of the patients also received prophylactic subcutaneous 200 µg of octreotide as an induction dose. Nasogastric (NG) tubes were routinely placed throughout the operation. Furthermore, two tubes were generally placed at the end of operation for drainage of fluid, i.e. a Jackson-Pratt drain (JP drain) near to the pancreatic stump remnant and another passive-drainage tube in the operation field. Simultaneously, postoperative pain was managed by an epidural anesthesia or patient-controlled analgesia (PCA) and all the patients were shifted to the intensive care unit (ICU) for a night. Besides, after surgery, some patients received a continuous intravenous infusion of octreotide at the rate of 0.25 mg/hr for 7 days with the help of a microinfusion pump on the random basis according to the surgeons preference.

Enhanced recovery after surgery (ERAS) protocol was used for postoperative management of all the patients, focusing on early mobilization and early nutrition intake^15^. Additionally, abdominal fluid drainage was monitored and if the amount of drainage fluid was < 10 ml after 24 hrs, the passive-drainage tube was withdrawn after an inspection with ultrasound to exclude any collection of fluid in the abdominal cavity. Moreover, the serum amylase level and drainage fluid amylase level (from JP drain) were examined after 3 days to rule out the presence of pancreatic fistula. In addition to this, at the time of follow-up Doppler ultrasound was used see the patency of splenic vessels (for the patients with spleen-preserving DP with preservation of splenic vessels) and to rule out any thrombus or stricture in vessels. Furthermore, all the data were documented prospectively in the hospital database.

### Operative Techniques

An operation was performed by 3 senior surgeons of our department. Moreover, the choice of surgical technique was decided by consultation between the surgeons and the patient party or according to underlying disease condition on preoperative radiological evaluation.

#### Surgical Procedure for laparoscopic DP

Laparoscopic DP was mostly carried out for benign and low-grade malignant tumors in the distal pancreas. The surgical techniques for laparoscopic DP has already been described in details in our previous papers^16, 17^. Generally, five trocars were inserted to perform DP. First of all, a 10 mm port incision was created just beneath the umbilicus for an observation hole. Additionally, pneumoperitoneum was created by the pressure of 13-15 mm of Hg. After creation of pneumoperitoneum, a trocar of 10 mm alone with 30° telescope was inserted into the abdominal cavity. Remaining, other four trocars were placed under the direct vision of the telescope above the umbilicus (two trocars, 12-mm and 5-mm on the right midclavicular line and remaining two 5-mm trocars on the left midclavicular line).

After ruling out any other abdominal pathology, metastasis, and any puncture to internal organs, abdominal surface of the pancreas was exposed by dissection of gastrocolic and gastrosplenic ligaments using a laparoscopic harmonic scalpel. Great care was taken to preserve the left gastroepiploic vessels and short gastric vessels. Further, the dissection was performed according to the surgeons preference, both superior-anterior approach^17^ and inferior-posterior approach^16^ are being used in our hospital for spleen-preserving DP with preservation of splenic vessels (Kimura technique^18^) by taking advantage of the avascular plain known as “the fusion fascia of Toldt”^16^. Warshaw technique^19^ was only performed for low-grade malignant tumor with suspected or known cases of tumor invading the splenic vessels. After obtaining adequate surgical margin and after sufficient mobilization of the pancreas, the pancreas was divided proximally approximately 2 cm far from the tumor with the help of Covidien Endo GIA Universal Straight 60-3.5 mm stapler. Additionally, in order to free distal pancreatic stump together with body and tail from splenic vessels, it was dissected dorsally with the help of an ultrasonic knife by pulling it to the left lateral side. Furthermore, to minimise the risk of POPF, in the recent years we routinely suture the pancreatic stump using polypropylene 3-0 intracorporeal interrupted sutures.

Nonetheless, splenectomy was performed in case of an inadequate blood supply and outflow obstruction of the spleen. Additionally, taking oncologic principle into consideration, splenectomy was also performed if the tumor lies in close proximity to the splenic hilum^20, 21^. At the end of the operation, the specimen was pulled out using a bag via an enlarged umbilical port-site incision, and was sent for histopathology. Besides, the texture of the pancreas was determined by the tactile feedback of the instrument and was reassured after being pulled out from the abdominal cavity. Lastly, the abdominal cavity was washed with warm water and a JP drain tube was placed close to the pancreatic stump and a passive-drainage tube in the operation field on the left side through 5 mm port-site incisions.

#### Surgical Procedure for Open DP

Open DP was carried out both for benign and malignant tumor in the distal pancreas. Open DP was performed with bilateral subcostal or upper midline incision. However, except the incision other techniques were somewhat similar to laparoscopic DP. Nonetheless, in most of the cases the transection of the pancreas was not done with Endo GIA stapler, the transection of pancreatic parenchyma was done using the surgical blade, and the main pancreatic duct on the remnant pancreatic stump was ligated using 4-0 or 5-0 polypropylene continuous suture whenever identified. Additionally, the remnant pancreatic stump was also sutured using 4-0 or 5-0 polypropylene continuous suture to avoid any leakage from the branch pancreatic duct. In all the cases of malignant tumor lymphadenectomy and the excision of the nodal tissues was performed along the common hepatic artery, the left gastric artery, the celiac axis, and along the superior mesenteric vein, including the peripancreatic lymph nodes. Additionally, extended resection along with resection of other visceral organs was performed in any cases of contiguous organ involvement.

### Definitions

The severity grading of surgical complications was determined as proposed by Clavien-Dendo classification^22^. Moreover, the postoperative complications like delayed gastric emptying (DGE),^23^ postpancreatectomy hemorrhage (PPH)^24^. and postoperative pancreatic fistula (POPF)^7^ were in accordance with the consensus definition of the International Study Group of pancreatic surgery (ISGPS). Additionally, postoperative mortality was defined as the death within 30 days after surgery or death during hospital the stay^25^.

### Management of the Pancreatic Fistula

All the cases of POPF were managed by drainage tube adjustment, extention of extubation time, adequate drainage, administration of octreotide, and antibiotic therapy. Additionally, relaparotomy was done for patients with Grade C POPF. However, there was no standard treatment protocol for the management of POPF.

### Statistical analysis

All the statistical analysis was performed using SPSS 16.0 (IBM Corp., Armonk, NY). Continuous data are reported as a mean±standard deviation (SD). Categorical data are reported as absolute numbers (n). The univariate analysis of risk factors for pancreatic fistula was performed by χ^2^ test, and the multivariate analysis was performed by multivariate logistic regression model (backward elimination method) to test the independent risk factors for pancreatic fistula. P<0.05 was considered statistically significant.

## Results

All 263 patients, including 124 males and 139 females underwent DP at the Li Hui Li Hospital and the Ningbo medical center, Ningbo between January 2011 and January 2020. The median age of the patients undergoing DP was 58 years (range 17-89 years). Of these 263 patients, 121 (46%) had malignant tumors and 142 (54%) had benign or low-grade malignant tumors (**Table 1)**. Among total patients, 213 patients underwent open surgery and 50 patients underwent laparoscopic surgery. The mean operation time was 221±90 minutes and mean blood loss was 375±215 ml. There were 165 cases of combined splenectomy, whereas spleen was preserved in 98 cases. However, endoscopy was not routinely performed at the time of follow-up, none of the patients suffered from gastric or esophageal variceal bleeding due to spleen-preserving DP. Multivisceral resections were carried out in 70 (26.6%) patients (4 patients had more than 2 combined organ resections) that includes- 32 partial gastrectomy, 8 adrenalectomy, 3 left nephrectomy, 18 partial hepatectomy, 13 partial small intestine or colon resection (**Table 2)**. Moreover, twenty one patients had extended pancreatic body and tail resection (i.e. the pancreas was cut to the right side of the portal vein).

**Table 1.** Primary lesions in 263 patients undergoing pancreatectomy

| **Primary lesion** | **Number of cases** | **Percentage ( % )** |
| --- | --- | --- |
| Pancreatic cancer | 85 | 32.3 |
| Pancreatic solid-pseudopapillary tumor | 9 | 3.4 |
| Other organ malignancy | 25 | 9.5 |
| Pancreatic trauma | 12 | 4.6 |
| Intraductal papillary mucinous neoplasm (IPMN) | 35 | 13.3 |
| Mucinous cystic neoplasm (MCN) | 23 | 8.7 |
| Pancreatic serous cystadenoma | 22 | 8.4 |
| Pancreatic cystadenocarcinoma | 5 | 1.9 |
| Primary pancreatic Non-Hodgkin's lymphoma | 4 | 1.5 |
| Pancreatic pseudocyst | 15 | 5.7 |
| Pancreatic abscess | 2 | 0.8 |
| Pancreatic neuroendocrine tumors | 12 | 4.6 |
| Pancreatic neuroendocrine carcinoma | 2 | 0.8 |
| Intrapancreatic Accessory Spleen | 1 | 0.4 |
| Chronic pancreatitis | 11 | 4.2 |

**Table 2.** Multivisceral Distal Pancreatectomy (n =70); Organs resected without Spleen (n=74)

| **Organ (Procedure)** | **Number** |
| --- | --- |
| Partial gastrectomy | 32 |
| Adrenalectomy | 8 |
| Left nephrectomy | 3 |
| Partial hepatectomy | 18 |
| Small intestine or colon resection | 13 |

Note: 4 patients had more than 2 combined organ resections.

### Management of Pancreatic Remnant

Mitigation techniques of the pancreatic remnant and resection margin was mainly done by two techniques in our series, i.e. 1. Manual closure using sutures 2. Closure using ENDO-GIA stapling. Manual closure using sutures was employed in 211 patients (80.2%) whereas ENDO-GIA stapling was used in 52 patients (19.7%). Of these, ligation of main pancreatic duct was performed in 174 patients (66.1%) overall. The incidence of CR-POPF was 23.1% in ENDO-GIA stapling and 18% in manual closure using sutures. However, the result was not statistically significant between the two.

### Pancreatic Fistula and Other Complications

The total postoperative complications developed in 38.4% (101/263) patients (i.e. one or more than one complications) that includes 50 cases of pancreatic fistula (19.0%), 10 cases of pulmonary infection (3.8%), 5 cases of abdominal bleeding (1.9%), 4 cases of cardiovascular complications (1.5%), 4 cases of chylous fistula (1.5%), 1 case complicated with biliary fistula, gastric fistula, severe abdominal infection, and renal failure in a trauma patient, which was managed after active treatment (0.4%), and 1 case of bile leakage in a patient with liver resection (0.4%) (**Table 3)**. Among the patients who suffered from POPF, there were 61 cases of biochemical leak, 48 cases (96.0%) of Grade B POPF, and 2 cases (4.0%) of Grade C POPF. The average time of hospital stay was 24.6±9.3 days in the patients with POPF and 19.8±7.3 days in the patients without POPF (P= 0.025). Similarly, the average time of hospital stay was 20.5 days±7.5 days in the patients with “biochemical leak" and 31.5 days ±9.2 days in the patients with CR-POPF (i.e Grade B and Grade C POPF). The result was statistically significant between the two groups (P= 0.038). Postoperative complications due CR-POPF occurred in 32 patients (64%) that includes, abdominal infection in 20 cases (40%), delayed PPH in 2 cases (4%), DGE in 5 cases (10%), and surgical site wound infection in 5 cases (10%). Fortunately, no postoperative mortality occurred in our series.

**Table 3.** Postoperative complications

| **Complication** | **N (%)** |
| --- | --- |
| Pancreatic fistula | 50 (19.0%) |
| Pulmonary infection | 10 (3.8%) |
| Abdominal bleeding | 5 (1.9%) |
| Cardiovascular complications | 4 (1.5%) |
| Chylous fistula | 4 (1.5%) |
| Biliary fistula, gastric fistula, severe abdominal infection, and renal failure in a trauma patient | 1 (0.4%) |
| Bile leakage in a patient with liver resection | 1 (0.4%) |

### Risk Factors for the Development of CR-POPF

Furthermore, all 263 patients were divided into CR-POPF group (n= 50) and non-CR-POPF group (n= 213) based on the occurrence of pancreatic fistula. The factors that might contribute in the development of pancreatic fistula are presented in **Table 4**. The univariate analysis of 18 factors showed that the patients with a malignant tumor, soft pancreas, and patient without ligation of the main pancreatic duct are more likely to develop pancreatic fistula. The incidence of CR-POPF in the patients with malignant tumor was 30/121 (24.8%) and 20/142 (14%) in the patients with benign disease or low-grade malignant tumors, P= 0.027. Similarly, the incidence of CR-POPF in the patients with soft pancreas was 25/88 (28.4%) and that of firm pancreas was 25/175 (14.2%), P= 0.006. Likewise, the incidence of CR-POPF in the patients without ligation of the main pancreatic duct was 26/89 (29.2%) and in the patients with ligation of the main pancreatic duct was 24/174 (13.8%). P= 0.003. However, univariate analysis demonstrated no significant relationship between CF-POPF and the following factors: age, BMI, smoking, ASA, combined multivisceral resection, splenectomy, pancreatic stump treatment, preoperative diabetes, intraoperative blood loss, use of somatostatin after surgery, preoperative albumin level, postoperative albumin level (3 days after surgery), surgical approach (open vs laparascopic), operation time, and pancreatic resection range. Only a significantly important association was demonstrated between CF-POPF and the following factors: pancreatic pathology (malignant tumor vs benign disease or low-grade malignant tumor: 24.8% vs 14%, P= 0.027), pancreas texture (soft vs firm: 28.4% vs 14.2%, P= 0.006), and ligation of the main pancreatic duct (No vs Yes: 29.2% vs 13.8%, P= 0.003).

**Table 4.** Univariate analysis of risk factors for postoperative pancreatic fistula after distal pancreatectomy (DP)

| Variables | Numb<br>er of<br>cases<br>n=<br>263 | Pancreatic<br>fistula<br>n= 50 | Non-pancreatic<br>fistula<br>n= 213 | $\chi^2$ | P value |
| --- | --- | --- | --- | --- | --- |
| Age |  |  |  |  |  |
| $\geq 70$ | 45 | 9 | 36 | 0.034 | 0.853 |
| $< 70$ | 218 | 41 | 177 | | |
| BMI ( kg / m <sup>2</sup> ) |  |  |  |  |  |
| $> 25$ | 84 | 11 | 73 | 2.806 | 0.094 |
| $\leq 25$ | 179 | 39 | 140 | | |
| Smoking |  |  |  |  |  |
| Yes | 67 | 13 | 54 | 0.009 | 0.925 |
| No | 196 | 37 | 159 |  |  |
| ASA |  |  |  |  |  |
| I | 114 | 21 | 93 | 0.046 | 0.831 |
| II-III | 149 | 29 | 120 |  |  |
| Indication for surgery |  |  |  |  |  |
| Benign disease or low-grade malignant tumors | 142 | 20 | 122 | 4.866 | 0.027 |
| Malignant tumors | 121 | 30 | 91 |  |  |
| Preoperative diabetes |  |  |  |  |  |
| Yes | 42 | 7 | 35 | 0.178 | 0.673 |
| No | 221 | 43 | 178 |  |  |
| Preoperative albumin level |  |  |  |  |  |
| ≥35 g/L | 227 | 45 | 182 | 0.711 | 0.399 |
| < 35 g/L | 36 | 5 | 31 |  |  |
| Surgical approach |  |  |  |  |  |
| Laparoscopic | 50 | 10 | 40 | 0.014 | 0.907 |
| Open | 213 | 40 | 173 |  |  |
| Operation time |  |  |  |  |  |
| ≥300 min | 54 | 10 | 44 | 0.011 | 0.918 |
| < 300 min | 209 | 40 | 169 |  |  |
| Pancreas texture |  |  |  |  |  |
| Soft | 88 | 25 | 63 | 7.586 | 0.006 |
| Hard | 175 | 25 | 150 |  |  |
| Pancreatic resection range |  |  |  |  |  |
| Left side of portal vein | 242 | 45 | 197 | 0.341 | 0.559 |
| Right side of portal vein | 21 | 5 | 16 |  |  |
| Splenectomy |  |  |  |  |  |
| Yes | 165 | 29 | 136 | 0.593 | 0.441 |
| No | 98 | 21 | 77 |  |  |
| Combined multivisceral resection |  |  |  |  |  |
| Yes | 70 | 13 | 57 | 0.012 | 0.913 |
| No | 193 | 37 | 156 |  |  |
| Ligation of main pancreatic duct |  |  |  |  |  |
| Yes | 174 | 24 | 150 | 9.094 | 0.003 |
| No | 89 | 26 | 63 |  |  |
| Pancreatic stump treatment |  |  |  |  |  |
| Suture | 211 | 38 | 173 | 0.696 | 0.404 |
| Endo-GIA Stapler | 52 | 12 | 40 |  |  |
| Intraoperative blood loss |  |  |  |  |  |
| ≥600 ml | 46 | 9 | 37 | 0.269 | 0.604 |
| < 600 ml | 217 | 41 | 176 |  |  |
| Use of somatostatin after surgery |  |  |  |  |  |
| Yes | 75 | 14 | 61 | 0.067 | 0.796 |
| No | 188 | 36 | 152 |  |  |
| Postoperative albumin level (3 days after surgery) |  |  |  |  |  |
| ≥35 g/L | 177 | 34 | 143 | 0.001 | 0.973 |
| < 35 g/L | 86 | 16 | 70 |  |  |

Note: BMI- Body Mass Index

Multivariate analysis was performed by multivariate logistic regression model (backward elimination method) for all 18 factors used in the univariate analysis. The results showed that the soft texture of the pancreas (OR= 2.381, P= 0.001) and the ligation of main pancreatic duct (OR= 0.388, P= 0.002) were an independent influencing factor for CR-POPF (**Table 5**). The ligation of main pancreatic duct was associated with lesser number of CR-POPF in the univariate analysis.

**Table 5.** Multivariate logistic regression analysis for postoperative pancreatic fistula after distal pancreatectomy (DP).

| Variables | $\beta$ | S.E. | Wald | *P* value | OR | 95% CI |
| --- | --- | --- | --- | --- | --- | --- |
| Pancreas texture<br>(Soft vs Hard) | -1.116 | 0.351 | 10.108 | 0.001 | 2.381 | 1.271-4.460 |
| Ligation of the<br>main pancreatic<br>duct (Yes vs<br>No) | -1.021 | 0.333 | 9.407 | 0.002 | 0.388 | 0.207-0.726 |

Note: $\beta$ = regression coefficient; S.E.= standard error of regression coefficient; Wald= Wald chi-square value; CI = confidence interval; OR = Odds ratio.

## Discussion

In this study, we have examined both mortality and morbidity related to DP, with particular aimed to POPF. Data from our study showed that DP can now be performed very safely without mortality. However, higher rate of the morbidity still remains the concern, which was close to 38.4% in our series. Particularly, CR-POPF was the most frequent complication that occured in 19% of our patients, the rate of CR-POPF in our series is similar to that reported in the literature^26-28^. Nonetheless, CR-POPF is the considerable contributor to major complications such as peripancreatic effusion, peripancreatic abscess, pseudocyst formation, or erosion and digestion of surrounding tissues, resulting in intra-abdominal hemorrhage, gastric emptying disorders, resulting in prolonged hospitalization time, increased hospitalization costs, affecting subsequent treatment following pancreatic resection^7-10^. Additionally, some patients may be readmitted after discharge due to the above complications. In our study, the length of hospital stay for CR-POPF group was significantly longer than that of non-CR-POPF group, with postoperative complications due CR-POPF occurred in 32 patients (64%) including bleeding.

On univariate analysis, CR-POPF occurred significantly at a higher rate in the soft pancreas (vs the hard pancreas) and on the other hand CR-POPF occurred significantly at a lower rate in the patients with benign disease or low-grade malignant tumor, and when intraoperative ligation of the main pancreatic duct was not done. No other factors were found to be related to an increased risk of CR-POPF. However, on multivariate analysis, only the texture of the pancreas and the ligation of main pancreatic duct were an independent influencing factor for CR-POPF.

Many other studies have revealed various preoperative, intraoperative, and postoperative, variables as the risk factors for the development of CR-POPF, i.e. age, intraoperative blood loss, soft texture of the pancreas, BMI, multivisceral resections, splenectomy, operation time, gland thickness, and fasting blood glucose level^26, 29-31^. However, most of these studies have been inconsistent with their findings with eachothers. The reasons for the inconsistent findings might be, retrospective nature of the studies, heterogeneous practices among the surgeons, and the consequences of a learning curve for CR-POPF occurrence and management in different centers. Thus, the relationship between different risk factors for the development of CR-POPF should be interpreted cautiously. Nonetheless, in most of the studies, soft texture of the pancreas has widely been recognized as the most significant risk factor for the development of CR-POPF^12, 26, 31^. In our series, 88 patients had soft pancreatic texture (CR-POPF 28.4%), and 175 patients had a hard pancreatic texture (CR-POPF 14.3%). Indeed, univariate analysis revealed there was significant statistical differences for the development of CR-POPF between the two groups (soft pancreatic texture vs hard pancreatic texture), P= 0.001, attributing that the patients with soft pancreatic texture were more prone to develop a CR-POPF after DP than patients with a hard pancreatic texture. Additionally, multivariate analysis implied that a soft pancreatic texture was an independent risk factor associated with CR-POPF (OR- 2.381 and 95% CI- 1.271- 4.460). The lower rate of CR-POPF in patients with hard pancreatic texture may be explained by pancreatic fibrosis resulting into the exocrinal dysfunction of the pancreas.

At present, the main mitigation strategies for pancreatic remnant to reduce risk of POPF includes, manual closure using sutures, closure using ENDO-GIA stapling, the use of fibrin glue to cover the pancreatic stump, coverage of the pancreatic stump with autologous tissue, the use of pancreatic stents, use of ultrasonic dissector, etc. ^10, 26^. Nonetheless, whether these mitigation strategies can reduce or prevent the occurrence of POPF is still debatable. There are several retrospective studies^26, 32^, randomized controlled trials (RCTs)^4, 11, 33^, and meta-analysis^34, 35^ evaluating these mitigation strategies and found no evidence that these techniques are able to prevent or reduce risks of developing CR-POPF. Results from our study suggests that the intraoperative ligation of the main pancreatic duct can reduce the incidence of POPF, this observation was consistent with previous studies^36-38^. In our study, the incidence of CR-POPF was 29.2% when there was no intraoperative ligation of the main pancreatic duct and 13.8% when there was intraoperative ligation of the main pancreatic duct. Nevertheless, elective ligation of the main pancreatic duct might be difficult sometimes, especially when the main pancreatic duct is too thin and it is difficult to be identified. To overcome such difficulties, we suggest sharp and careful transection of the pancreatic body or tail, where the main pancreatic duct can easily be identified in most of the cases. However, we should also acknowledge that, ligation of only main pancreatic duct is not an ultimate solution for POPF, the opening of the small branch ducts on the margin of the pancreatic remnant may also cause POPF. Because of the contractile resistance of the sphincter of Oddi, the pressure of the main pancreatic duct increases, which results in the formation of POPF due to the opening of the accessory branched pancreatic ducts on the pancreatic remnant. Thus, manual closure using sutures on the margin of the remnant pancreatic stump may be necessary. However, POPF can easily occur in the soft pancreas due to cutting and tearing of pancreatic tissue by sutures. Furthermore, if the suture is densely placed on the pancreatic remnant, it may cause ischemic necrosis of the tissue in the remnant pancreatic stump. Similarly, if the suture is placed too loose, it will cause POPF due to the incomplete suturing of the pancreatic stump. Thus, surgeons must take these factors into consideration while suturing the main pancreatic duct and the remnant pancreatic stump. However, some authors believe that the ligation of the main pancreatic duct does not affect the occurrence of pancreatic fistula^26, 28^. It has been reported that preoperative endoscopic pancreatic stent implantation can effectively reduce the pressure of pancreatic exocrine ducts, thereby reducing the occurrence of pancreatic fistula^39^. Additionally, more recently in a study by Ecker et. al reported that the use of epidural analgesia was associated with significantly fewer incidence of POPF, probably because of it is able to reduce the sphincter of Oddi pressure^26^. On the other hand, some authors believe that POPF can effectively be reduced by anastomosis of pancreatic stump to stomach (pancreatico-gastrostomy)^40^ or to jejunum (pancreatico-jejunostomy)^41^ after DP^42^. However, the accuracy of these additional operations to prevent POPF remains to be further confirmed, but these additional surgical procedures undoubtedly will increase the complexity of the operation and prolong the time of the operation. In other words, this may increase the possibility of other postoperative complications. For the internal drainage of pancreatic stump, the authors believe that, if preoperative imaging or intraoperative exploration reveals obstruction of the proximal pancreatic duct, the pancreatic stump should be anastomosed with jejunum or the posterior gastric wall to drain the pancreatic juice, which may prevent POPF caused by the proximal pancreatic duct pressure.

ENDO-GIA stapling is a common method in DP for closure of of pancreatic stump, especially for laparascopic surgery. It has advantages that it can save operation time and can be performed easily compared to the transection of pancreatic parenchyma using the surgical blade. However, there are some unfavorable factors, such as inadequate ligation of main pancreatic duct and tension at the edge of pancreatic stump, which aggravate local ischemia and necrosis of the pancreatic stump. In our series, the incidence of CR-POPF in ENDO-GIA stapling group was 23.1% and 18% in stuture group. However, there was no significant difference between both the groups. The reason we speculate for this is that, in the recent years we routinely suture the pancreatic stump using polypropylene 3-0 intracorporeal interrupted sutures after ENDO-GIA stapling. Thus, this might have influenced the incidence of CR-POPF in ENDO-GIA stapling group. Therefore, we believe that manual suture still remains the mainstream method for the treatment of pancreatic stump after DP.

Our study has several limitations that need to be emphasised. Firstly, this study is a retrospective nature and thus, subject to biases. Likewise, the data included in this study is over a long period of time (2011–2020) and may have different surgical techniques and POPF mitigation strategies depending upon individual surgeon preference. Similarly, there might be a potential mis-grading of patients with biochemical leakage before the updated definition of ISGPS 2016. Secondly, some clinical data are not sufficient like we couldn’t collect proper data for pancreatic thickness, where different studies has outlined it as an independent risk factor for CR-POPF^43, 44^. Thirdly, the effects of a learning curve on POPF occurrence and management of POPF cannot be excluded. Finally, our Electronic Medical Record System might not have the record of complications that were managed in the local hospitals. However, on the other hand our study is still of great importance, as it includes large size of the cases from a single center. Moreover, we have analysed most of the clinically relevant variables that might have an effect on the occurrence of POPF in both open and laparascopic DP.

## Conclusion

Pancreatic fistula was the most common surgical complication after DP. The texture of pancreas and ligation of main pancreatic duct can influence the incidence of CR-POPF. No other factors like, age, BMI, smoking, ASA, combined multivisceral resection, splenectomy, pancreatic stump treatment, preoperative diabetes, intraoperative blood loss, use of somatostatin after surgery, preoperative albumin level, postoperative albumin level (3 days after surgery), surgical approach (open vs laparascopic), operation time, and pancreatic resection range had an influence on development of CR-POPF after DP.

## Data Availability

All the data supporting the results were shown in the paper and are available from the corresponding author upon request.

## Conflicts of Interest

The authors declare no competing interests.

## Authors’ Contributions

Gao Qing Wang and Dipesh Kumar Yadav have equally contributed in study design, data collection, data analysis, and preperation of the manuscript.

## Study design

Gao Qing Wang, Dipesh Kumar Yadav, and Cai De Lu

## Data collection

Gao Qing Wang, Dipesh Kumar Yadav, Wei Jiang, and Yong Fei Hua

## Data analysis

Gao Qing Wang, Dipesh Kumar Yadav

## Preperation of the manuscript

Gao Qing Wang, Dipesh Kumar Yadav

## Final draft review

Gao Qing Wang, Dipesh Kumar Yadav, Wei Jiang, Yong Fei Hua, and Cai De Lu

## Acknowledgments

This study was funded by

1. The Ningbo Natural Science Foundation of China (2019A610214).
2. Ningbo Health Branding Subject Fund (PPXK2018-03).
3. The Scientific Innovation Team Project of Ningbo (2013B82010).

## Notes

### Competing Interest Statement

The authors have declared no competing interest.

## References

1. Kawaida H, Kono H, Watanabe M, et al. Risk factors of postoperative pancreatic fistula after distal pancreatectomy using a triple-row stapler. Surgery today 2018; 48: 95–100. 2017/06/11. DOI: 10.1007/s00595-017-1554-2.

2. Sugimoto M, Gotohda N, Kato Y, et al. Risk factor analysis and prevention of postoperative pancreatic fistula after distal pancreatectomy with stapler use. Journal of hepato-biliary-pancreatic sciences 2013; 20: 538–544. 2013/02/23. DOI: 10.1007/s00534-013-0596-0.

3. Okano K, Oshima M, Kakinoki K, et al. Pancreatic thickness as a predictive factor for postoperative pancreatic fistula after distal pancreatectomy using an endopath stapler. Surgery today 2013; 43: 141–147. 2012/07/12. DOI: 10.1007/s00595-012-0235-4.

4. Diener MK, Seiler CM, Rossion I, et al. Efficacy of stapler versus hand-sewn closure after distal pancreatectomy (DISPACT): a randomised, controlled multicentre trial. Lancet (London, England) 2011; 377: 1514–1522. 2011/05/03. DOI: 10.1016/s0140-6736(11)60237-7.

5. Eguchi H, Nagano H, Tanemura M, et al. A thick pancreas is a risk factor for pancreatic fistula after a distal pancreatectomy: selection of the closure technique according to the thickness. Digestive surgery 2011; 28: 50–56. 2011/02/05. DOI: 10.1159/000322406.

6. Seeliger H, Christians S, Angele MK, et al. Risk factors for surgical complications in distal pancreatectomy. American journal of surgery 2010; 200: 311–317. 2010/04/13. DOI: 10.1016/j.amjsurg.2009.10.022.

7. Bassi C, Marchegiani G, Dervenis C, et al. The 2016 update of the International Study Group (ISGPS) definition and grading of postoperative pancreatic fistula: 11 Years After. Surgery 2017; 161: 584–591.2017/01/04. DOI: 10.1016/j.surg.2016.11.014.

8. Lee MKt, Lewis RS, Strasberg SM, et al. Defining the post-operative morbidity index for distal pancreatectomy. HPB: the official journal of the International Hepato Pancreato Biliary Association 2014; 16: 915–923. 2014/06/17. DOI: 10.1111/hpb.12293.

9. Pratt WB, Maithel SK, Vanounou T, et al. Clinical and economic validation of the International Study Group of Pancreatic Fistula (ISGPF) classification scheme. Annals of surgery 2007; 245: 443–451. 2007/04/17. DOI: 10.1097/01.sla.0000251708.70219.d2.

10. Nahm CB, Connor SJ, Samra JS, et al. Postoperative pancreatic fistula: a review of traditional and emerging concepts. Clinical and experimental gastroenterology 2018; 11: 105–118. 2018/03/29. DOI: 10.2147/ceg.s120217.

11. Miyasaka Y, Mori Y, Nakata K, et al. Attempts to prevent postoperative pancreatic fistula after distal pancreatectomy. Surgery today 2017; 47: 416–424. 2016/06/22. DOI: 10.1007/s00595-016-1367-8.

12. Callery MP, Pratt WB, Kent TS, et al. A prospectively validated clinical risk score accurately predicts pancreatic fistula after pancreatoduodenectomy. Journal of the American College of Surgeons 2013; 216: 1–14. 2012/11/06. DOI: 10.1016/j.jamcollsurg.2012.09.002.

13. Malik AY and Foster C. The revised Declaration of Helsinki: cosmetic or real change? J R Soc Med 2016; 109: 184–189. DOI: 10.1177/0141076816643332.

14. Doyle DJ and Garmon EH. American Society of Anesthesiologists Classification (ASA Class). StatPearls. Treasure Island (FL): StatPearls Publishing LLC., 2019.

15. Joliat GR, Labgaa I, Petermann D, et al. Cost-benefit analysis of an enhanced recovery protocol for pancreaticoduodenectomy. The British journal of surgery 2015; 102: 1676–1683. 2015/10/23. DOI: 10.1002/bjs.9957.

16. Hua YF, Yadav DK, Bai X, et al. Laparoscopic Spleen-Preserving Distal Pancreatectomy (LSPDP) with Preservation of Splenic Vessels: An Inferior-Posterior Approach. Gastroenterology research and practice 2018; 2018: 1683719–1683719. DOI: 10.1155/2018/1683719.

17. Huang J, Yadav DK, Xiong C, et al. Laparoscopic Spleen-Preserving Distal Pancreatectomy (LSPDP) versus Open Spleen-Preserving Distal Pancreatectomy (OSPDP): A Comparative Study. Canadian Journal of Gastroenterology and Hepatology 2019; 2019: 7. DOI: 10.1155/2019/9367868.

18. Kimura W, Yano M, Sugawara S, et al. Spleen-preserving distal pancreatectomy with conservation of the splenic artery and vein: techniques and its significance. Journal of hepato-biliary-pancreatic sciences 2010; 17: 813–823. 2009/12/22. DOI: 10.1007/s00534-009-0250-z.

19. Warshaw AL. Conservation of the spleen with distal pancreatectomy. Archives of surgery (Chicago, 111:1960) 1988; 123: 550–553.1988/05/01. DOI: 10.1001/archsurg.1988.01400290032004.

20. Strasberg SM, Drebin JA and Linehan D. Radical antegrade modular pancreatosplenectomy. Surgery 2003; 133: 521–527. 2003/05/30. DOI: 10.1067/msy.2003.146.

21. Christein JD, Kendrick ML, Iqbal CW, et al. Distal pancreatectomy for resectable adenocarcinoma of the body and tail of the pancreas. Journal of gastrointestinal surgery: official journal of the Society for Surgery of the Alimentary Tract 2005; 9: 922–927. 2005/09/03. DOI: 10.1016/j.gassur.2005.04.008.

22. Clavien PA, Barkun J, de Oliveira ML, et al. The Clavien-Dindo classification of surgical complications: five-year experience. Annals of surgery 2009; 250:187–196. 2009/07/30. DOI: 10.1097/SLA.0b013e3181b13ca2.

23. Wente MN, Bassi C, Dervenis C, et al. Delayed gastric emptying (DGE) after pancreatic surgery: a suggested definition by the International Study Group of Pancreatic Surgery (ISGPS). Surgery 2007; 142: 761–768. 2007/11/06. DOI: 10.1016/j.surg.2007.05.005.

24. Wente MN, Veit JA, Bassi C, et al. Postpancreatectomy hemorrhage (PPH): an International Study Group of Pancreatic Surgery (ISGPS) definition. Surgery 2007; 142: 20–25. 2007/07/17. DOI: 10.1016/j.surg.2007.02.001.

25. Jacobs JP, Mavroudis C, Jacobs ML, et al. What is operative mortality? Defining death in a surgical registry database: a report of the STS Congenital Database Taskforce and the Joint EACTS-STS Congenital Database Committee. The Annals of thoracic surgery 2006; 81: 1937–1941. 2006/04/25. DOI: 10.1016/j.athoracsur.2005.11.063.

26. Ecker BL, McMillan MT, Allegrini V, et al. Risk Factors and Mitigation Strategies for Pancreatic Fistula After Distal Pancreatectomy: Analysis of 2026 Resections From the International, Multi-institutional Distal Pancreatectomy Study Group. Annals of surgery 2019; 269: 143–149. 2017/09/01. DOI: 10.1097/sla.0000000000002491.

27. Dokmak S, Fteriche FS, Meniconi RL, et al. Pancreatic fistula following laparoscopic distal pancreatectomy is probably unrelated to the stapler size but to the drainage modality and significantly decreased with a small suction drain. Langenbeck’s archives of surgery 2019; 404: 203–212. 2019/02/11. DOI: 10.1007/s00423-019-01756-3.

28. Nathan H, Cameron JL, Goodwin CR, et al. Risk factors for pancreatic leak after distal pancreatectomy. Annals of surgery 2009; 250: 277–281. 2009/07/30. DOI: 10.1097/SLA.0b013e3181ae34be.

29. Kleeff J, Diener MK, Z’Graggen K, et al. Distal pancreatectomy: risk factors for surgical failure in 302 consecutive cases. Annals of surgery 2007; 245: 573–582. 2007/04/07. DOI: 10.1097/01.sla.0000251438.43135.fb.

30. Ke Z, Cui J, Hu N, et al. Risk factors for postoperative pancreatic fistula: Analysis of 170 consecutive cases of pancreaticoduodenectomy based on the updated ISGPS classification and grading system. Medicine (Baltimore) 2018; 97: e12151. 2018/09/02. DOI: 10.1097/md.0000000000012151.

31. Xia T, Zhou JY, Mou YP, et al. Risk factors for postoperative pancreatic fistula after laparoscopic distal pancreatectomy using stapler closure technique from one single surgeon. PLoS One 2017; 12: e0172857. 2017/02/25. DOI: 10.1371/journal.pone.0172857.

32. Fahy BN, Frey CF, Ho HS, et al. Morbidity, mortality, and technical factors of distal pancreatectomy. American journal of surgery 2002; 183: 237–241. 2002/04/12. DOI: 10.1016/s0002-9610(02)00790-0.

33. Carter Tl, Fong ZV, Hyslop T, et al. A dual-institution randomized controlled trial of remnant closure after distal pancreatectomy: does the addition of a falciform patch and fibrin glue improve outcomes? Journal of gastrointestinal surgery: official journal of the Society for Surgery of the Alimentary Tract 2013; 17: 102–109. 2012/07/17. DOI: 10.1007/sll605-012-1963-x.

34. Probst P, Huttner FJ, Klaiber U, et al. Stapler versus scalpel resection followed by hand-sewn closure of the pancreatic remnant for distal pancreatectomy. The Cochrane database of systematic reviews 2015: Cd008688. 2015/11/07. DOI: 10.1002/14651858.CD008688.pub2.

35. Knaebel HP, Diener MK, Wente MN, et al. Systematic review and meta-analysis of technique for closure of the pancreatic remnant after distal pancreatectomy. The British journal of surgery 2005; 92: 539–546. 2005/04/27. DOI: 10.1002/bjs.5000.

36. Bilimoria MM, Cormier JN, Mun Y, et al. Pancreatic leak after left pancreatectomy is reduced following main pancreatic duct ligation. The British journal of surgery 2003; 90: 190–196. 2003/01/30. DOI: 10.1002/bjs.4032.

37. Pannegeon V, Pessaux P, Sauvanet A, et al. Pancreatic fistula after distal pancreatectomy: predictive risk factors and value of conservative treatment. Archives of surgery (Chicago, Ill: 1960) 2006; 141: 1071–1076; discussion 1076. 2006/11/23. DOI: 10.1001/archsurg.141.11.1071.

38. Distler M, Kersting S, Rückert F, et al. Chronic pancreatitis of the pancreatic remnant is an independent risk factor for pancreatic fistula after distal pancreatectomy. BMC Surg 2014; 14: 54–54. DOI: 10.1186/1471-2482-14-54.

39. Abe N, Sugiyama M, Suzuki Y, et al. Preoperative endoscopic pancreatic stenting for prophylaxis of pancreatic fistula development after distal pancreatectomy. American journal of surgery 2006; 191: 198–200. 2006/01/31. DOI: 10.1016/j.amjsurg.2005.07.036.

40. Yanagimoto H, Satoi S, Toyokawa H, et al. Pancreaticogastrostomy following distal pancreatectomy prevents pancreatic fistula-related complications. Journal of hepato-biliary-pancreatic sciences 2014; 21: 473–478. 2013/12/18. DOI: 10.1002/jhbp.59.

41. Kawai M, Hirono S, Okada K, et al. Randomized Controlled Trial of Pancreaticojejunostomy versus Stapler Closure of the Pancreatic Stump During Distal Pancreatectomy to Reduce Pancreatic Fistula. Annals of surgery 2016; 264:180–187. 2015/10/17. DOI: 10.1097/sla.0000000000001395.

42. Tieftrunk E, Demir IE, Schorn S, et al. Pancreatic stump closure techniques and pancreatic fistula formation after distal pancreatectomy: Meta-analysis and single-center experience. PLOS ONE 2018; 13: e0197553. DOI: 10.1371/journal.pone.0197553.

43. Kim H, Jang J-Y, Son D, et al. Optimal stapler cartridge selection according to the thickness of the pancreas in distal pancreatectomy. Medicine 2016; 95: e4441–e4441. DOI: 10.1097/MD.0000000000004441.

44. Kawai M, Tani M, Okada K, et al. Stump closure of a thick pancreas using stapler closure increases pancreatic fistula after distal pancreatectomy. American journal of surgery 2013; 206: 352–359. 2013/06/29. DOI: 10.1016/j.amjsurg.2012.11.023.

